# Co-occurrence of SARS-CoV-2 and Respiratory Pathogens in the Frail Elderly

**DOI:** 10.1101/2020.06.24.20138941

**Authors:** Alan Wolfe, David Baunoch, Dakun Wang, Ryan Gnewuch, Xinhua Zhao, Thomas Halverson, Patrick Cacdac, Shuguang Huang, Trisha Lauterbach, Natalie Luke

**Affiliations:** Department of Microbiology and Immunology, Loyola University Chicago, Maywood, IL; Stat4ward, 711 Parkview Dr., Gibsonia, PA 15044; Pathnostics, 17661 Cowan, Irvine, CA 92614; Capstone Laboratory, 8601 Dunwoody Pl #444, Sandy Springs, GA 30350

**Keywords:** SARS-CoV-2, COVID19, Respiratory Pathogens, Nursing Homes, Assisted Living Facilities, Long Term Care

## Abstract

**Background:** Elderly SARS-CoV-2 patients are associated with higher hospitalization and mortality. Co-infection is critical in the severity of respiratory diseases. It is largely understudied for SARS-CoV-2.

**Methods:** Between March 24^th^ and April 27^th^, 2020, nasopharyngeal and oropharyngeal swabs from 3,348 patients from nursing homes and assisted living facilities in 22 states in the US were tested by Capstone Healthcare for SARS-CoV-2, 24 other respiratory viruses, and 8 respiratory bacteria. Total nucleic acid was extracted with MagMAX™ Viral/Pathogen Ultra nucleic acid isolation kit. SARS-Co-V-2 was detected with the CDC 2019-novel coronavirus (2019-nCoV) diagnostic panel. Total nucleic acid was pre-amplified before analysis for other respiratory pathogens with Taqman OpenArray™ Respiratory Tract Microbiota Plate.

**Results:** Patients’ mean age was 76.9 years. SARS-CoV-2 was detected in 1,413 patients (42.2%). Among them, 1,082 (76.6%) and 737 (43.7%) patients were detected with at least one bacterium or another virus, respectively. SARS-CoV-2-positive patients were more likely to have bacterial co-occurrences (76.6%) than SARS-CoV-2-negative patients (70.0%) (p<0.0001). The most common co-occurring bacteria were *Staphylococcus aureus* and *Klebsiella pneumonia*, detected in 55.8% and 40.1% SARS-CoV-2-positive patients, respectively. *Staphylococcus aureus* was associated with SARS-CoV-2, with higher detection rates in SARS-CoV-2-positive patients (55.8%) than SARS-CoV-2-negative patients (46.2%) (p<0.0001). Human herpes virus 6 (HHV6) also was common and associated with SARS-CoV-2, with higher detection rates in SARS-CoV-2-positive patients (26.6%) than SARS-CoV-2-negative patients (19.1%) (p<0.0001).

**Conclusions:** SARS-CoV-2-positive patients are more likely to be positive for certain respiratory bacteria and viruses. This observation may help explain high hospitalization and mortality rates in older patients.

## Introduction

As of May 12, 2020, the SARS-CoV-2 pandemic has resulted in more than 3.5 million confirmed cases and 250,000 deaths (1). In the US alone, there are more than 1.3 million confirmed cases, with an overall cumulative hospitalization rate of 50.3 per 100,000 (2). In the US, the highest rates of hospitalization is in people 65 years and older (162.2 per 100,000), who contribute 80% of the 78,000 deaths (3).

In the ongoing battle against SARS-CoV-2, much effort has focused on screening and testing effective anti-viral drugs and developing vaccines. In contrast, co-infection is largely understudied. This is problematic, as co-infection has proven to be critical in the severity of other respiratory diseases and significantly impacts patient clinical management, infection control, and antimicrobial stewardship programs.

Secondary bacterial pneumonia, a common form of co-infection, was a predominant cause of death in the 1918-1919 and the 1957-1958 influenza pandemics (4). Bacterial co-infection also was reported in approximately 20% of the 2009 H1N1 influenza patients, and was associated with poor outcome (5,6). Thus, it is reasonable to suspect significant levels of co-infection in the current pandemic. The level of co-infection is particularly important as it is reported that broad-spectrum empirical antibiotics are frequently prescribed (72%) in SARS-CoV-2 patients despite a paucity of evidence of bacterial co-infection (7). A better understanding of co-infection levels in patients with SARS-CoV-2 will facilitate better evaluation of patient prognosis and allow development of more effective clinical management plans (8).

Co-infection with coronavirus, especially with its three most prominent members, SARS-CoV-1, MERS and SARS-CoV-2, has been reported only infrequently, due in part to the novel nature of this viral group (7). For SARS-CoV-2, co-infection with bacteria, other respiratory viruses, and/or fungi has been described, with a mean co-infection rate of approximately 8% across all studies (7). For example, Zhou et al. reported 15% secondary lung infection that existed in 50% of all fatal cases (9). Chen et al. reported 4% co-infection of bacteria and fungi with SARS-CoV-2 (10). However, most studies were case reports or based on scattered available clinical information, and not designed to systematically evaluate the prevalence or the characteristics of co-infections. Two studies were exceptions. Lin et al. used multiplex reverse transcription–polymerase chain reaction (RT-PCR) to simultaneously detect SARS-CoV-2 and 14 other respiratory viruses in sputum, nasal or throat swabs from 186 highly suspected COVID-19 cases between January 20^th^ and February 1^st^, 2020 obtained from multiple sites in the city of Shenzhen, China. Among the 92 SARS-CoV-2-postive patients, co-infection with other viruses was only identified in 6 patients (3.2%), whereas 18 out of the 94 SARS-CoV-2-negatvie patients (9.7%) had co-infections. Bacterial co-infection was not tested (11). In a more recent study, Kim et al. analyzed 1217 nasopharyngeal swabs from 1206 symptomatic patients (with cough, fever, and/or dyspnea) received between March 3^rd^ and March 27^th^, 2020 from multiple sites in northern California. These authors also used RT-PCR technology to test for SARS-CoV-2 and a panel of other pathogens, which included 12 other respiratory viruses or viral groups, as well as 2 bacteria species. A total of 116 of the 1217 specimens (9.5%) were identified as positive for SARS-CoV-They reported no statistically significant difference in co-infection rates with other respiratory pathogens in patients positive [24 of 116 (20.7%)] or negative [294 of 1101 (26.7%)] for SARS-Co-2 (difference 6.0%, 95% CI, −2.3% to 14.3%) (12).

Nursing homes and assisted living facilities are at high risk for SARS-CoV-2. Their residents are older adults living in close quarters; most have underlying chronic medical conditions. In this study, we used RT-PCR technology to investigate the co-occurrence of SARS-Co-V-2 with a large panel of other respiratory pathogens in 3,348 patients from nursing homes and assisted living facilities in the US.

## Methods

### Patients and specimens

Capstone Healthcare tested a total of 3,348 adult patients from 156 nursing homes and assisted living facilities from 22 states in the US for both SARS-CoV-2 and other respiratory pathogens. Specimens included in the study were collected via nasopharyngeal and oropharyngeal swabs between March 24^th^ and April 27^th^, 2020, transported to Capstone Healthcare in molecular transport medium, and processed as described below. The study was exempted from Institutional Review Board (IRB) review by Western IRB.

### Total nucleic acid extraction

Total nucleic acid was extracted in the BSL-3 laboratory at Capstone Healthcare with MagMAX™ Viral/Pathogen Ultra nucleic acid isolation kit, according to the manufacturer’s instruction (Cat. No. A42356, Thermo Fisher, Waltham, MA).

### Detection of SARS-Co-V-2

Detection of SARS-Co-V-2 was performed with the CDC 2019-Novel Coronavirus (2019-nCoV) Real-Time RT-PCR Diagnostic panel according to CDC’s instruction (Cat. No. 2019-nCoVEUA-01, CDC, Atlanta GA). The one-step RT-PCR was performed with the total extracted nucleic acid as the template on a QuantStudio™12K Flex Real-time PCR system. SARS-CoV-2 status was determined based on RT-PCR results from 2019-nCoV markers N1 and N2, and RNase P, an extraction control, according to the CDC’s instruction.

### Detection of other respiratory pathogen

The extracted nucleic acid was pre-amplified on a Veriti™ Thermal Cycler before analysis for presence of other respiratory pathogens on the QuantStudio™12K Flex Real-time PCR system with Taqman OpenArray™ Respiratory Tract Microbiota Plate (Cat. No. A41237, Thermo Fisher, Waltham, MA), according to the manufacturer’s instructions outlined in the application guide for Respiratory Tract Microbiota Profiling Experiments (Pub. Num. MAN0017952). The OpenArray™ Respiratory Tract Microbiota Plates included 24 respiratory viruses and 8 bacteria measured in triplicate for each specimen. The viruses include influenza A/B, influenza A/H1-2009, influenza A/H3, human herpesvirus 3/4/5/6, parainfluenza virus type 1/2/3/4, adenovirus, rhinovirus, human bocavirus, coronavirus HKU, coronavirus OC43, coronavirus NL63, coronavirus 229E, human metapneumovirus, respiratory syncytial virus A/B, human enterovirus, human enterovirus D68. The bacteria included *Haemophilus influenzae, Legionella pneumophila, Klebsiella pneumonia, Staphylococcus aureus, Mycoplasma pneumonia, Bordetella bronchiseptica/parapertussis/ Bordetella pertussis, Streptococcus pneumonia*, and *Chlamydophila pneumonia*.

### Statistical analysis

Patient and facility characteristics were described by frequency and percentages. SARS-CoV-2 detection rate among the full sample, and the co-occurrence of respiratory bacteria and other respiratory viruses among SARS-CoV-2-positive patients were calculated and compared by patient characteristics using chi-square test, separately.

The overall associations between SARS-CoV-2 with respiratory bacteria and other respiratory viruses were reported by crosstab and chi-square test. The three-way detection rate of SARS-CoV-2, respiratory bacteria, and other respiratory viruses were depicted in a bar chart. The infection rate of each individual respiratory bacterium and other respiratory virus was calculated overall and compared by SARS-CoV-2-positive/negative status. Multivariate logistic regression model with robust variance at facility level to account for clustering of patients within the same facility was used to assess whether SARS-CoV-2 was independently associated with the co-occurrence of respiratory bacteria. Lastly, sensitivity analyses were run by excluding the patients with multiple tests to assess the robustness of the study findings. The analyses were performed using SAS version 9.4 and StataSE 16.

## Results

### Patient characteristics

Capstone Healthcare tested a total of 3,586 nasopharyngeal and oropharyngeal specimens from 3,348 adult patients; 3,115 had one test, 228 had 2 tests, and 5 had 3 tests. Among those 233 patients with multiple tests, only 23 had results changed, including 8 patients changed from positive to negative and 15 changed from negative to positive. For the patients with multiple tests, results from the first test were included in the analysis with the following exception: for the 15 patients whose SARS-CoV-2 results changed from negative to positive, the second test set, associated with the positive SARS-CoV-2 result, was included in the analysis.

Table 1 provides detailed characteristics of the study subjects. All patients were from either nursing homes (55.8%) or assisted living facilities (44.2%). The mean age was 76.9 years old, with 78.6% subjects being 65 years or older. There were 2,367 (70.7%) females and 981 (29.3%) males. Patients were mainly from three states, FL (22.5%), GA (25.1%), and NJ (24.0%).

**Table 1.** Patient characteristics and SARS-CoV-2 positive rates. Patient gender, age, facility type and location by state, and SARS-CoV-2 positivity status are summarized in table 1. * Indicates the states with less than 10 patients enrolled in the study.

| N=3,348 patients | Sample Distribution | N=1,413<br>SARS-CoV-2-positive patients |  |
| --- | --- | --- | --- |
|  |  | N (%) | P value |
| **Gender, N (%)** |  |  | 0.54 |
| Female | 2367 (70.7) | 991 (41.9) |  |
| Male | 981 (29.3) | 422 (43.0) |  |
| **Age, mean ± SD**<br>**(min, max)** | 76.9 ± 16.6<br>(18, 106) | | <0.0001 |
| 18-39 | 154 (4.6) | 25 (16.2) |  |
| 40-64 | 562 (16.8) | 175 (31.1) |  |
| 65-84 | 1330 (39.7) | 627 (47.1) |  |
| 85+ | 1302 (38.9) | 586 (45.0) |  |
| **Facility Type** |  |  | <0.0001 |
| Assisted Living | 1479 (44.2) | 440 (29.7) |  |
| Nursing Home | 1869 (55.8) | 973 (52.1) |  |
| **Facility State** |  |  | <0.0001 |
| CO | 20 (0.6) | 16 (80.0) |  |
| FL | 752 (22.5) | 102 (13.6) |  |
| GA | 841 (25.1) | 334 (39.7) |  |
| IN | 243 (7.3) | 82 (33.7) |  |
| MA | 236 (5.8) | 175 (77.1) |  |
| NJ | 803 (24.0) | 550 (68.5) |  |
| NY | 242 (7.2) | 60 (24.8) |  |
| OH | 13 (0.4) | 5 (38.5) |  |
| PA | 95 (2.8) | 58 (61.1) |  |
| TX | 80 (2.4) | 23 (28.8) |  |
| Other* | 32 (1.0) | 8 (25.0) |  |
| Do Not Know | 15 (0.4) | 11 (73.3) |  |

\*States with less than 10 patients (AL, AR, DC, DE, IL, KS, KY, LA, ME, MI, RI, VA)

### SARS-CoV-2 infection status

Of the 3,348 patients, 1,413 (42.2%) tested positive for SARS-CoV-2, with no significant difference between males and females (p=0.54) (Table 1). In contrast, older patients (≥ 65) tested positive more frequently than younger ones (p<0.0001). Test results also differed by facility type with more positive results from nursing homes than from assisted living facilities (<0.0001). They also differed by location (state) of the facility (<0.0001).

### Co-occurrence of SARS-CoV-2 and other pathogens

Table 2 lists the co-occurrence of respiratory bacteria and other respiratory viruses with SARS-CoV-2, stratified by gender, age, facility type and facility location (by state). In total, of the 1,413 SARS-CoV-2-positive patients, 1,082 (76.6%) and 734 (52.2%) were detected with at least one respiratory bacterium or other respiratory virus, respectively. Patient age (p=0.005) and facility type (<0.0001) were associated with co-occurrence of SARS-CoV-2 with respiratory bacteria, but not with other respiratory viruses. Facility location was more strongly associated with respiratory bacteria (p<0.0001) than other respiratory viruses (p=0.019).

**Table 2.**
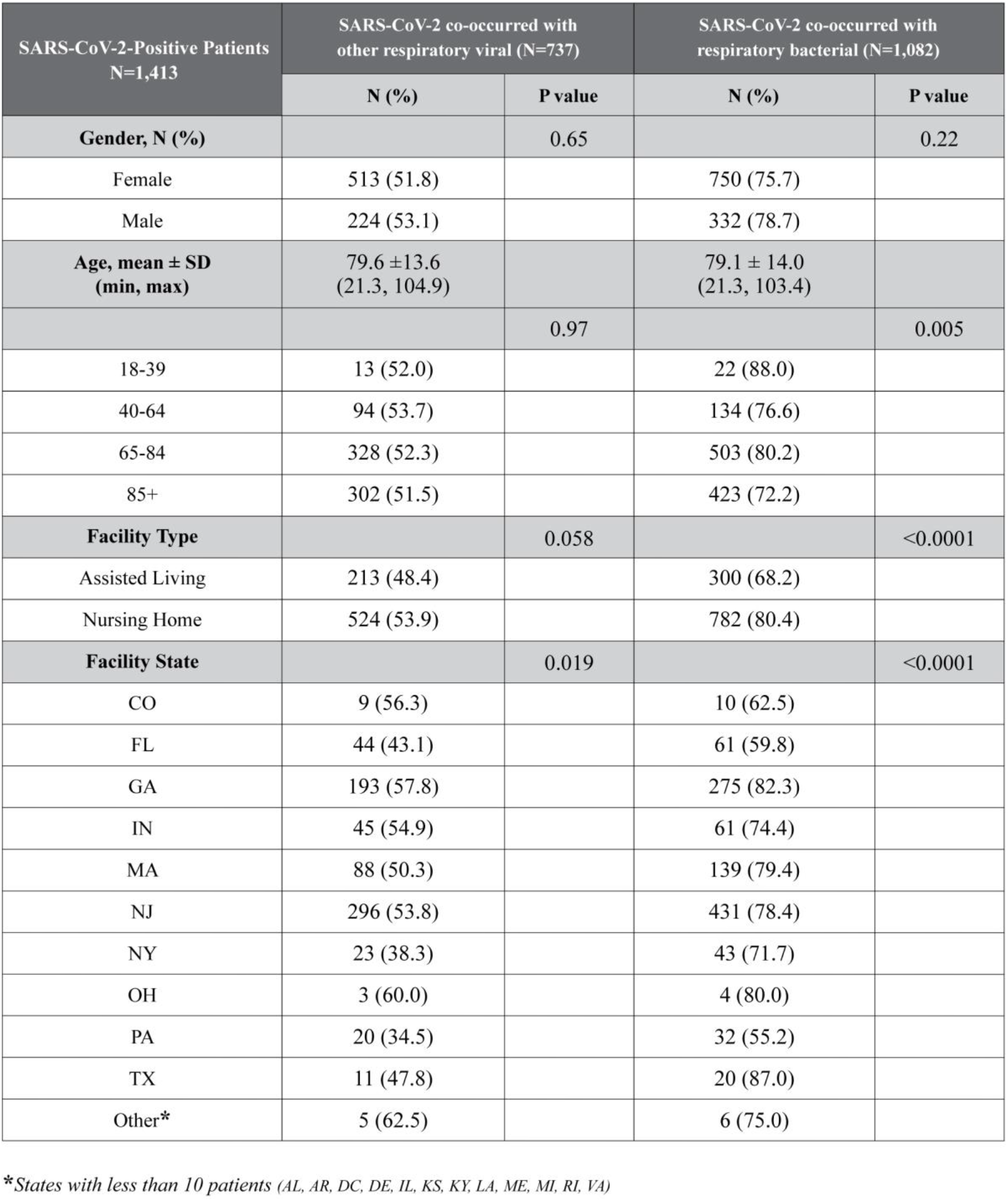
Co-occurrences of SARS-CoV-2 with other respiratory viruses and bacteria. Co-occurrences of SARS-CoV-2 with other respiratory viruses and bacteria stratified with patient gender, age, facility type and location by state are summarized in table 2. * Indicates states with less than 10 patients enrolled in the study.

| SARS-CoV-2-Positive Patients<br>N=1,413 | SARS-CoV-2 co-occurred with<br>other respiratory viral (N=737) |  | SARS-CoV-2 co-occurred with<br>respiratory bacterial (N=1,082) |  |
| --- | --- | --- | --- | --- |
|  | N (%) | P value | N (%) | P value |
| **Gender, N (%)** |  | 0.65 |  | 0.22 |
| Female | 513 (51.8) |  | 750 (75.7) |  |
| Male | 224 (53.1) |  | 332 (78.7) |  |
| **Age, mean ± SD**<br>**(min, max)** | 79.6 ± 13.6<br>(21.3, 104.9) | | 79.1 ± 14.0<br>(21.3, 103.4) | |
|  |  | 0.97 |  | 0.005 |
| 18-39 | 13 (52.0) |  | 22 (88.0) |  |
| 40-64 | 94 (53.7) |  | 134 (76.6) |  |
| 65-84 | 328 (52.3) |  | 503 (80.2) |  |
| 85+ | 302 (51.5) |  | 423 (72.2) |  |
| **Facility Type** |  | 0.058 |  | <0.0001 |
| Assisted Living | 213 (48.4) |  | 300 (68.2) |  |
| Nursing Home | 524 (53.9) |  | 782 (80.4) |  |
| **Facility State** |  | 0.019 |  | <0.0001 |
| CO | 9 (56.3) |  | 10 (62.5) |  |
| FL | 44 (43.1) |  | 61 (59.8) |  |
| GA | 193 (57.8) |  | 275 (82.3) |  |
| IN | 45 (54.9) |  | 61 (74.4) |  |
| MA | 88 (50.3) |  | 139 (79.4) |  |
| NJ | 296 (53.8) |  | 431 (78.4) |  |
| NY | 23 (38.3) |  | 43 (71.7) |  |
| OH | 3 (60.0) |  | 4 (80.0) |  |
| PA | 20 (34.5) |  | 32 (55.2) |  |
| TX | 11 (47.8) |  | 20 (87.0) |  |
| Other* | 5 (62.5) |  | 6 (75.0) |  |

\*States with less than 10 patients (AL, AR, DC, DE, IL, KS, KY, LA, ME, MI, RI, VA)

Table 3 compares SARS-CoV-2-positive and –negative patients. SARS-CoV-2-positive patients were more likely to test positive for at least one respiratory bacterium (1,082/1,413 [76.6%]) than SARS-CoV-2-negative patients (1,354/1,935 [70.0%]) (p<0.0001). No association was observed for other respiratory viruses (737/1,413 [52.2%] vs. 949/1,935 [49.0%] for SARS-CoV-2-positive and -negative patients, respectively [p=0.075]). Multivariate analysis showed that SARS-CoV-2 status was an independent factor associated with bacterial co-infection, regardless of patient age, gender, facility type and state location (odds ratio=1.26, 95% confidence interval: 1.04-1.54; p=0.02). More than 600 patients tested positive for SARS-CoV-2, at least one respiratory bacterium, and at least one other respiratory virus, which translates to 42.7% (603/1,413) co-occurrence with both a respiratory bacterium and another respiratory virus in SARS-CoV-2-positive patients (Figure 1).

**Table 3.** Cross tab of the test results of SARS-CoV-2 with other respiratory viruses and bacteria.

| N<br>Column percent | Co-occurrence | SARS-CoV-2 |  |  | P value |
| --- | --- | --- | --- | --- | --- |
|  |  | Negative<br>N=1,935<br>57.8% | Positive<br>N=1,413<br>42.2% | Total |  |
| Other<br>Respiratory<br>Viruses | Negative | 986<br>50.96% | 676<br>47.84% | 1662<br>49.64% | P=0.075 |
|  | Positive | 949<br>49.04% | 737<br>52.16% | 1686<br>50.36% |  |
| Respiratory<br>Bacteria | Negative | 581<br>30.03% | 331<br>23.43% | 912<br>27.24% | P<0.0001 |
|  | Positive | 1354<br>69.97% | 1082<br>76.57% | 2346<br>72.76% |  |

**Figure 1.**
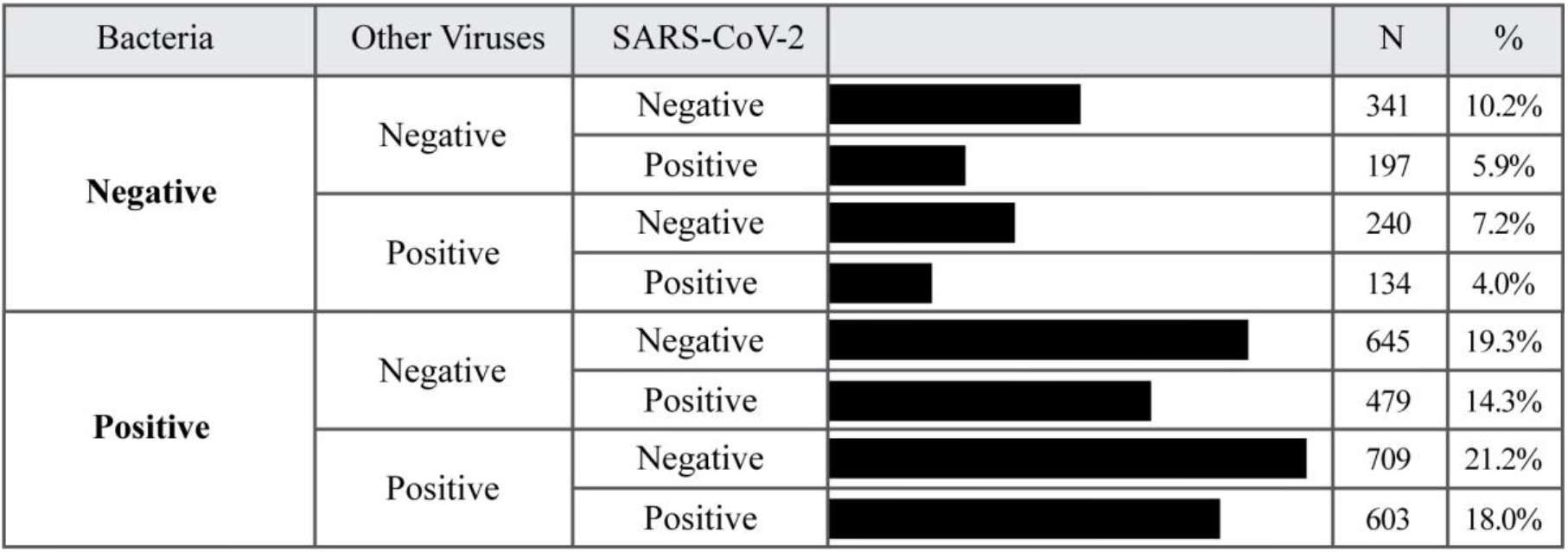
Frequencies of co-occurrence of SARS-CoV-2, other respiratory viruses, and respiratory bacteria

Table 4 shows the co-occurrence data for SARS-CoV-2, respiratory bacteria, and other respiratory viruses. All 24 other respiratory viruses and 8 bacteria were detected in at least one patient, with two exceptions: *Chlamydophila pneumonia* and *Legionella pneumophila*. The most common co-occurring bacterial species were *Staphylococcus aureus, Klebsiella pneumoniae* and *Haemophilus influenza*, detected in 55.8%, 40.1% and 17.2% of SARS-CoV-2-positive patients, respectively. More importantly, *S. aureus* was associated with infection of SARS-CoV-2, with detection rates of 55.8% and 46.2% (p<0.0001) in SARS-CoV-2-positive and -negative patients, respectively. *K. pneumoniae* was also associated with SARS-CoV-2. None of the other bacteria were associated with SARS-CoV-2. The most common co-occurring viruses were human herpesvirus 4 (HHV4 - Epstein-Barr Virus) and human herpesvirus 6 (HHV6). The majority of the viral detections (22/24, 91.7%) were not associated with detection of SARS-CoV-2, with the exception of HHV6 and rhinovirus. The detection rates of HHV6 in SARS-CoV-2-positive and SARS-CoV-2-negative patients were 26.6% vs. 19.1% (p<0.0001), respectively. A small number of patients (0.1% to 0.4%) tested positive for both influenza (A/B or other subtypes) and SARS-CoV-2. Co-occurrence of other coronaviruses with SARS-CoV-2, such as human coronavirus OC43 (0.1%), HKU1 (0.5%), 229E (0.0%), and NL63 (0.5%), were also low (Supplementary Table 1).

**Table 4.** The detection rates of individual other organisms, overall and by SARS-CoV-2 status. SARS-CoV-2 and a total of 32 other respiratory viruses and bacteria were tested in the study. The 32 organisms include 24 other respiratory viruses and 8 respiratory bacteria. Table 4 only lists the 8 organisms (4 other respiratory viruses and 4 respiratory bacteria) with prevalence of 1% or above. Detection and co-occurrence information for the organisms with less than 1% prevalence are listed in Supplementary Table 1.

| Organism (32) | Total N=3,348 | SARS-CoV-2 |  |  |
| --- | --- | --- | --- | --- |
|  |  | Negative<br>N=1,935 | Positive<br>N=1,413 | P value |
| **Other Respiratory virus (24)\*** | 1686 (50.4) | 949 (49.0) | 737 (52.2) | 0.080 |
| Human herpesvirus 4 (HHV4 - Epstein-Barr) | 1166 (34.8) | 669 (34.6) | 497 (35.2) | 0.74 |
| Human herpesvirus 6 (HHV6) | 745 (22.3) | 369 (19.1) | 376 (26.6) | **<0.0001** |
| Human Rhinovirus | 54 (1.6) | 40 (2.1) | 14 (1.0) | **0.017** |
| Human herpesvirus 5 (HHV5 - Cytomegalovirus) | 48 (1.4) | 25 (1.3) | 23 (1.6) | 0.46 |
| **Respiratory Bacteria (8)\*** | 2436 (72.8) | 1354 (70.0) | 1082 (76.6) | **<0.0001** |
| *Staphylococcus aureus* | 1683 (50.3) | 894 (46.2) | 789 (55.8) | **<0.0001** |
| *Klebsiella pneumoniae* | 1257 (37.5) | 691 (35.7) | 566 (40.1) | **0.011** |
| *Haemophilus influenzae* | 558 (16.7) | 315 (16.3) | 243 (17.2) | 0.48 |
| *Streptococcus pneumoniae* | 146 (4.4) | 90 (4.7) | 56 (4.0) | 0.35 |

\*Only organisms (8) with prevalence of 1% or above, 4 other respiratory viruses and 4 respiratory bacteria, were presented in the table, among the 32 tested in our study. The details for other organisms were listed in Supplementary Table 1.

The results were very similar from the sensitivity analysis by excluding those 233 patients with multiple tests.

## Discussion

The SARS-CoV-2 pandemic is a major challenge to healthcare systems. Proper diagnosis and management of elderly patients are critical due to their high hospitalization and mortality rates. Experiences from past influenza pandemics demonstrated the important role of bacterial co-infection in the severity of respiratory diseases (4-6). Our observation that SARS-CoV-2-positive patients were more likely also to be positive for certain respiratory bacteria and viruses may help explain high hospitalization and mortality rates in older patients.

The rates of co-occurrence in our study were considerably higher than those from the Lin et al. and Kim et al. studies. This difference may be due to several reasons. First, more pathogens were included in this study. A total of 24 respiratory viruses and 8 respiratory bacteria were tested in this study, compared with 12 viruses in the former study, and 12 viruses and viral groups, and 2 bacteria (*Chlamydia pneumoniae* and *Mycoplasma pneumoniae*) in the latter study. In this study, *Chlamydia pneumoniae* was not detected in any patients and *Mycoplasma pneumoniae* was only detected in 3 patients, and none of these patients were SARS-CoV-2-positive. Second, the patients included in our study were much older, with mean age of 76.9 years and only 21.4% younger than 65 years. The mean age in the Kim et al. study was 45.1 years. Likewise, 78% of patients were <65 years in the Lin et al. study. Age, suboptimal medical condition and immune-compromised status, congregated living environment and other factors may make these elderly patients more susceptible to respiratory infections. Third, all the patients in our study were from nursing homes and assisted living facilities. Fourth, the specimens were collected primarily in April for our study, but January and March for the other two studies (10,11).

The high bacterial co-occurrence rates in our study may support the clinical use of antibiotics in the management of elderly patients with SARS-CoV-2. The majority of study patients were positive for at least one respiratory bacterium, and the detection rate was higher in SARS-CoV-2– positive patients. The most commonly detected bacterial species in SARS-CoV-2-postive patients was *S. aureus*, which also was positively associated with SARS-CoV-2 infection. This Gram-positive bacterium is commonly carried on the skin or in the nose of healthy people (13). It also is a recognized cause of influenza-associated community-acquired pneumonia (14, 15). It was a relatively minor cause of 1918 influenza fatalities, but was predominant in the 1957–1958 influenza pandemic (4,14,16,17). It also was one of the most common bacteria co-infected with the 2009 H1N1 influenza virus, in addition to *Streptococcus pneumoniae* and *Streptococcus pyogenes* (6). In the current study, *S. aureus* was detected in 56.2% of SARS-CoV-2-positive patients, compared with 46.2% of SARS-CoV-2-negative patients (10% difference, p<0.0001). Clinically, some *S. aureus* strains are antibiotic-resistant, such as methicillin-resistant *S. aureus* (MRSA), and vancomycin-resistant *S. aureus* (VRSA). These findings may help increase the awareness of the potential presence of hard-to-treat bacterial infections, such as MRSA or VRSA in patients with SARS-CoV-2. The other two commonly detected bacteria were the Gram-negatives *Klebsiella pneumonia* and *Haemophilus influenza*, but the former was weakly associated, whereas the latter was not associated.

The high rate of *S. aureus* co-occurrence with SARS-CoV-2 may support the clinical use of broad-spectrum antibiotics in SARS-CoV-2 patients. Results from this study also provide pertinent information for healthcare facilities to adapt their antibiotic stewardship programs, in response to the pandemic. Whether co-occurrences with other pathogens result in clinically relevant co-infections, and whether they indeed contribute to the severity and the outcome of SARS-CoV-2 are important and intriguing clinical questions that warrant further investigations.

Results from the current study also confirmed previous reports that suggest the importance of testing patients with respiratory infectious diseases with both SARS-CoV-2 and other respiratory viruses, and that identification of other respiratory virus cannot exclude SARS-CoV-2, and vise versa. The co-occurrence rates of influenza viruses were low in both SARS-CoV-2-positive and - negative patients, presumably due to the fact that the specimen collection period occurred at the end of the flu season and lockdown policies were commonly in place due to the pandemic. The two most commonly detected viruses identified in our study were HHV4 (Epstein-Barr virus) and HHV6. Both viruses have been identified as risk factors for cognitive deterioration and progression to Alzheimer’s disease in elderly persons (18). Many people become infected with HHV4 and HHV6 in childhood. The viruses then became latent, during which time they may replicate in the salivary glands and be secreted in saliva without inducing any obvious pathology. In immune-compromised persons, reactivation of virus from latency or reinfection may occur (19,20). However, in this study, only HHV6, not HHV4, was associated with SARS-CoV-2 positivity, which may be due to selective reactivation of HHV6 in SARS-CoV-2-positive patients. HHV6 is known to have broad tropism (21). This virus may be present, whereas HHV4 may not, in pneumocytes and/or angiotensin-converting enzyme 2 (ACE2)-positive cells that are targeted by the SARS-CoV-2 virus. The exact underlying mechanisms of the selective association with SARS-CoV-2 of HHV6, but not HHV4, and the clinical significance of this co-occurring virus in disease severity or response to potential treatment in patients with SARS-CoV-2 should be investigated further.

In summary, this co-occurrence study of SARS-CoV-2 and 32 other respiratory pathogens in a large cohort of patients from multiple nursing homes and assisted living facilities in 22 states in the US demonstrated that SARS-CoV-2-positive patients are more likely to be positive for certain respiratory bacteria and viruses. This observation may help explain high hospitalization and mortality rates in older patients.

## Data Availability

Data is available upon request by writing to

## Acknowledgements

This work was supported by Pathnostics.

## Author Contributions

**Concept and design:** Baunoch, Wolf

**Acquisition, analysis, or interpretation of data:** Lauterbach, Gnewuch

**Drafting of the manuscript:** Wang

**Critical revision of the manuscript for important intellectual content:** Wolfe, Baunoch, Luke, Huang, Zhao, Halverson, Cacdac

## Statistical analysis

Zhao, Huang

**Administrative, technical, or material support:** Wang, Luke

**Supervision:** Luke

